# Twelve-year changes in protein profiles in patients with and without gastric bypass surgery

**DOI:** 10.1101/2020.08.13.20173666

**Authors:** Noha A. Yousri, Rudolf Engelke, Hina Sarwath, Rodrick D. McKinlay, Steven C. Simper, Ted D. Adams, Frank Schmidt, Karsten Suhre, Steven C. Hunt

## Abstract

Gastric bypass surgery results in long-term weight loss due to re-routing of the gastro-intestinal anatomy and dietary intake alterations. Studies have examined protein change during rapid weight loss (up to 1 year post-surgery), but whether protein changes are maintained long-term after weight stabilization is unknown. To identify proteins and pathways involved with the long-term beneficial effects of weight loss, abundances of 1297 blood-circulating proteins were measured at baseline, 2 and 12 years after Roux-en-Y gastric bypass surgery. Protein changes were compared between 234 surgery and 144 non-surgery subjects with severe obesity, with discovery and replication subgroups. Seventy-one protein changes were associated with 12-year BMI changes and 58 (7 unique) with surgical status. Protein changes, including ApoM, were most strongly associated with long-term changes in lipids (HDL-C and triglycerides). Inflammation, adipogenesis, cellular signaling, and complement pathways were implicated. Short-term improvements in protein levels were maintained long-term, even after some weight regain.

## Introduction

Bariatric surgery results in significant, sustained weight loss in most patients. The surgery also generally improves most cardiovascular risk factors, including blood pressure, sleep apnea, diabetes, dyslipidemia, fatty liver disease, levels of inflammation and health-related quality of life^1-5^. Total mortality and cardiovascular mortality are reduced by 40% and 56%, respectively^6,7^. In addition, bariatric surgery reduces the incidence (24-33%)^8,9^ and mortality of cancer (46%)^8^. Where voluntary, non-surgical weight loss is difficult to maintain over the long term^10^, surgical weight loss is generally maintained long-term, and as such, may provide an efficient way of identifying proteins and protein pathways possibly contributing to improved health outcomes induced by bariatric surgery. Discovery of these weight-related proteins and pathways may eventually be useful to improve weight loss and health in patients who are obese but do not elect or qualify for bariatric surgery.

A number of recent short-term studies have suggested particular plasma proteins that significantly change after bariatric surgery^11-14^. However, since most of these studies have examined protein changes only up to one year, (i.e. the nadir of maximum weight loss^15^), it is unknown which protein changes are durable in the long term. The effects of weight regain after maximum weight loss on protein levels is also unknown. Long-term assessment is important because acute post-surgery changes in many proteins may be due to the invasive nature of the surgery itself. The dramatic first year post-surgery weight-loss and the attendant changes in diet would be expected to affect many related physiological systems. However, once weight has stabilized and modest weight regain progresses, the underlying short-term benefits of the protein changes may disappear if these proteins return to near pre-surgical levels. Since gastric bypass surgery has been shown to improve many clinical risk factors for cardiovascular disease 10 to 15 years after surgery, identification of proteins that remain changed in the long-term may help identify biological pathways responsible for the reduction in disease.

We performed a SomaLogic (SomaLogic, Boulder, CO) aptamer-based proteomics study of 1297 proteins from 378 subjects with severe obesity who were followed for 12-years, with exams at baseline (exam 1), two years (exam 2), six years (exam 3) and 12 years (exam 4)^4^. Two hundred and thirty-four of the 378 subjects had gastric bypass surgery after the baseline exam. All protein measurements were performed prior to data analysis in discovery and replication subsets to provide within-study validation of results.

The hypotheses tested were: 1) Protein changes between exam 1 and exam 4 would be significantly associated and replicated with 12-year change in BMI (surgery and non-surgery groups combined); 2) Protein changes between exam 1 and exam 4 would be significantly different between the gastric bypass surgery and non-surgery groups; 3) Some of the protein change differences found from testing hypothesis 2 would be associated with surgery group status even after adjustment for change in BMI (suggesting the effects of surgery include factors other than just change in BMI); 4) The superset of protein changes identified in hypotheses 1-3 would show similar but opposite in direction changes per unit BMI during the main weight-loss period from baseline to 2 years compared to the changes per unit BMI during the remaining 10-year follow-up period where weight was either maintained or partially regained; and 5) The identified protein changes would be associated with clinical risk factors for diabetes, dyslipidemia, hypertension, liver disease, and cardiovascular disease.

## Results

There were 203 subjects in the discovery set and the 175 subjects in the replication set who had protein measurements at the baseline (exam 1) and at 12-year follow-up (exam 4; Table 1). Of the 234 surgery subjects in the two batches, 204 also had proteomics measured at 2-year (exam 2). Proteins in the non-surgery subjects were not measured at two years. A 6-year follow-up exam was performed (exam 3), but no proteins were measured. Characteristics of the subjects are shown in Table 1.

**Table 1.** Discovery and replication subject subset characteristics and winsorized variable means and standard deviations

| Variable |  | Discovery |  |  | Replication |  |  |
| --- | --- | --- | --- | --- | --- | --- | --- |
| Groups |  | Surgery<br>(n=137) | Non-Surgery<br>(n=66) | All<br>(n=203) | Surgery<br>(n=97) | Non-Surgery<br>(n=78) | All<br>(n=175) |
| **Gender (M)** |  | 20% | 12% | 17% | 12% | 11% | 12% |
| **Age (T1, y)** |  | 46.4 ± 10.5 | 47.1 ± 11 | 46.6 ± 10.9 | 40.1 ± 10 | 43.9 ± 12.3 | 41.8 ± 11.2 |
| **BMI (T1, kg/m<sup>2</sup>)** |  | 46.4 ± 7.1 | 44.6 ± 6.4 | 45.8 ± 6.9 | 46.1 ± 6.4 | 45.3 ± 7.5 | 45.8 ± 6.9 |
| **BMI (T2, kg/m<sup>2</sup>)**<br>**(n=204)** |  | 29.7 ± 5.5<br>(n=137) | NA | NA | 34.3 ± 9.1<br>(n=67) | NA | NA |
| **BMI (T4, kg/m<sup>2</sup>)** |  | 34.6 ± 8 | 44.8 ± 7.5 | 37.9 ± 9.1 | 35.0 ± 8.8 | 45.0 ± 8.9 | 39.6 ± 10.2 |
| **Changes in Clinical Variables (exam 4 – exam 1): Mean ± Standard Deviation, N** |  |  |  |  |  |  |  |
| FFM (kg) | M±SD | -13.4±6.7 | -2.4±6.5 | -9.9±8.4 | -13.5±7.0 | -4.9±7.4 | -9.8±8.4 |
|  | N | 127 | 59 | 186 | 91 | 70 | 161 |
| FM (kg) | M±SD | -22.2±11.5 | -2.1±10.3 | -15.8±14.5 | -21.0±11.6 | -1.2±10.1 | -12.4±14.7 |
|  | N | 127 | 59 | 186 | 91 | 70 | 161 |
| BMI<br>(kg/m<sup>2</sup>) | M±SD | -11.5±5.4 | -0.2±5.6 | -7.8±7.6 | -11.0±5.5 | -0.2±5.3 | -6.2±7.6 |
|  | N | 137 | 66 | 203 | 97 | 78 | 175 |
| REE<br>(kcal) | M±SD | -550±238 | -301±215 | -478±257 | -569±260 | -325±249 | -452±282 |
|  | N | 79 | 32 | 111 | 50 | 46 | 96 |
| AST<br>(U/l) | M±SD | -0.5±8.1 | -2.4±8.5 | -1.1±8.3 | -0.5±10.0 | -4.0±7.8 | -2.1±9.2 |
|  | N | 136 | 66 | 202 | 96 | 77 | 173 |
| ALT<br>(U/l) | M±SD | -5.0±11.3 | -2.7±13.1 | -4.3±11.9 | -4.4±15.0 | -5.4±12.8 | -4.8±14.0 |
|  | N | 137 | 66 | 203 | 97 | 77 | 174 |
| Uric acid<br>(mg/dl) | M±SD | -0.7±1.2 | 0.1±1.3 | -0.4±1.3 | -0.8±1.3 | -0.2±1.2 | -0.6±1.3 |
|  | N | 137 | 65 | 202 | 96 | 77 | 173 |
| SBP<br>(mmHg) | M±SD | -4.6±19.1 | 9.3±21.5 | -0.1±20.9 | -2.0±21.5 | 8.8±20.3 | 2.8±21.6 |
|  | N | 137 | 66 | 203 | 97 | 78 | 175 |
| DBP<br>(mmHg) | M±SD | -1.4±13.9 | 5.5±14.0 | 0.8±14.3 | 1.1±14.9 | 7.0±14.4 | 3.7±14.9 |
|  | N | 137 | 66 | 203 | 97 | 78 | 175 |
| Glucose<br>(mg/dl) | M±SD | -13.3±20.7 | -2.5±23.9 | -9.8±22.3 | -7.5±14.0 | -1.9±25.9 | -5.0±20.3 |
|  | N | 137 | 66 | 203 | 97 | 78 | 175 |
| Insulin<br>(μU/ml) | M±SD | -10.8±11.8 | -6.7±13.9 | -9.5±12.6 | -11.3±13.0 | -6.2±12.8 | -9.1±13.1 |
|  | N | 137 | 66 | 203 | 97 | 78 | 175 |
| HOMA-IR | M±SD | -3.1±3.4 | -2.0±4.1 | -2.8±3.7 | -2.9±3.5 | -1.5±3.9 | -2.3±3.7 |
|  | N | 137 | 66 | 203 | 97 | 78 | 175 |
| HOMA-B | M±SD | -71±147 | -69±177 | -70.0±157 | -100±179 | -38±166 | -73±175 |
|  | N | 133 | 63 | 196 | 96 | 76 | 172 |
| HbA1c<br>(%) | M±SD | 0.01±0.93 | 0.25±1.00 | 0.09±0.96 | 0.07±0.76 | 0.44±1.22 | 0.23±1.01 |
|  | N | 136 | 66 | 202 | 97 | 78 | 175 |
| TG<br>(mg/dl) | M±SD | -70.6±59.8 | -24.8±56.8 | -55.7±62.5 | -73.0±67.3 | -33.9±69.7 | -55.6±70.9 |
|  | N | 137 | 66 | 203 | 97 | 78 | 175 |
| LDL-C<br>(mg/dl) | M±SD | -2.9±29.5 | 26.4±29.7 | 6.6±32.5 | -3.4±30.6 | 27.8±28.4 | 10.5±33.4 |
|  | N | 137 | 66 | 203 | 97 | 78 | 175 |
| HDL-C<br>(mg/dl) | M±SD | 13.7±12.6 | 2.2±9.4 | 10.0±12.8 | 15.6±13.8 | 2.3±9.0 | 9.7±13.6 |
|  | N | 137 | 66 | 203 | 97 | 78 | 175 |
| FRS (%) | M±SD | -2.3 ± 3 | -0.8 ± 2 | -1.8 ± 3 | -1.0 ± 2 | -0.8 ± 2 | -0.9 ± 2 |
|  | N | 137 | 66 | 203 | 97 | 78 | 175 |
| DMINC | % | 0 | 0 | 0 | 3 | 21 | 24 |
|  | N | 91 | 41 | 132 | 88 | 51 | 139 |
| DMREM | % | 23 | 0 | 23 | 4 | 5 | 9 |
|  | N | 46 | 25 | 71 | 8 | 25 | 33 |

T1: baseline exam; T2: exam at 2 years; T4: exam at 12 years; BMI: body mass index; NA: not measured at T2; FFM: fat-free mass; FM (fat mass); REE: resting energy expenditure; AST (aspartate aminotransferase); ALT (alanine aminotransferase); SBP: systolic blood pressure; DBP: diastolic blood pressure; HOMA-IR: homeostatic assessment of insulin resistance; HOMA-B: homeostatic assessment of insulin secretion; HbA1c: hemoglobin A1c; TG: triglycerides; LDL-C: low-density lipoprotein cholesterol; HDL-C: high-density lipoprotein cholesterol; FRS: Framingham risk score for 10-year cardiovascular disease incidence. DMINC: Diabetes incidence; DMREM: Diabetes remission. Only 204 of the 234 subjects who had gastric bypass surgery had available T2 samples for proteomic measurements.

### Seventy-one proteins were associated with changes in BMI after 12-years

In the discovery set, 12-year changes in levels of 99 proteins were associated with changes in BMI after adjustment for age, gender, and baseline BMI, with correction for multiple comparisons (p<0.05/1297). Of the 99 protein changes associated with change in BMI, 71 were replicated (p<0.05/99; Table 2).. Sixty proteins increased as BMI decreased over the 12 years, while 11 proteins decreased (Table 2, Figure 1). Proteins that decreased with decreasing BMI were leptin (LEP), C-reactive protein (CRP), growth hormone receptor (GHR), afamin (AFM), serum amyloid P-component (APCS), reticulon-4 receptor (RTN4R), C5a anaphylatoxin (C5.1), heparin cofactor 2 (SERPIND1), pigment epithelium-derived factor (SERPINF1), ficolin-2 (FCN2), and myeloperoxidase (MPO). Of the 60 proteins that increased with decreasing BMI, the most strongly associated were IGFBP2, WFIKKN2, NCAM1, APOM, and ADIPOQ.

**Table 2.**
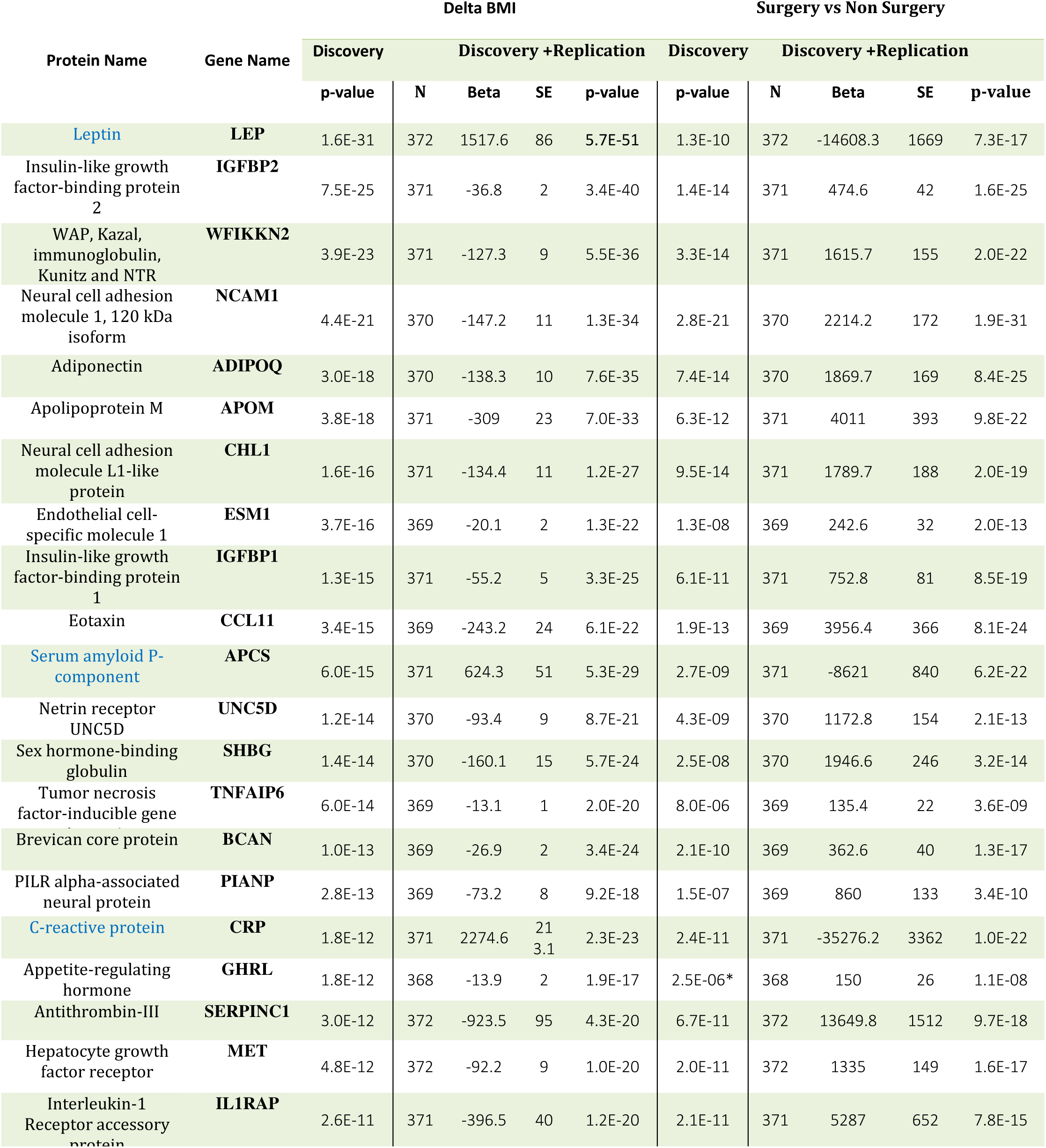
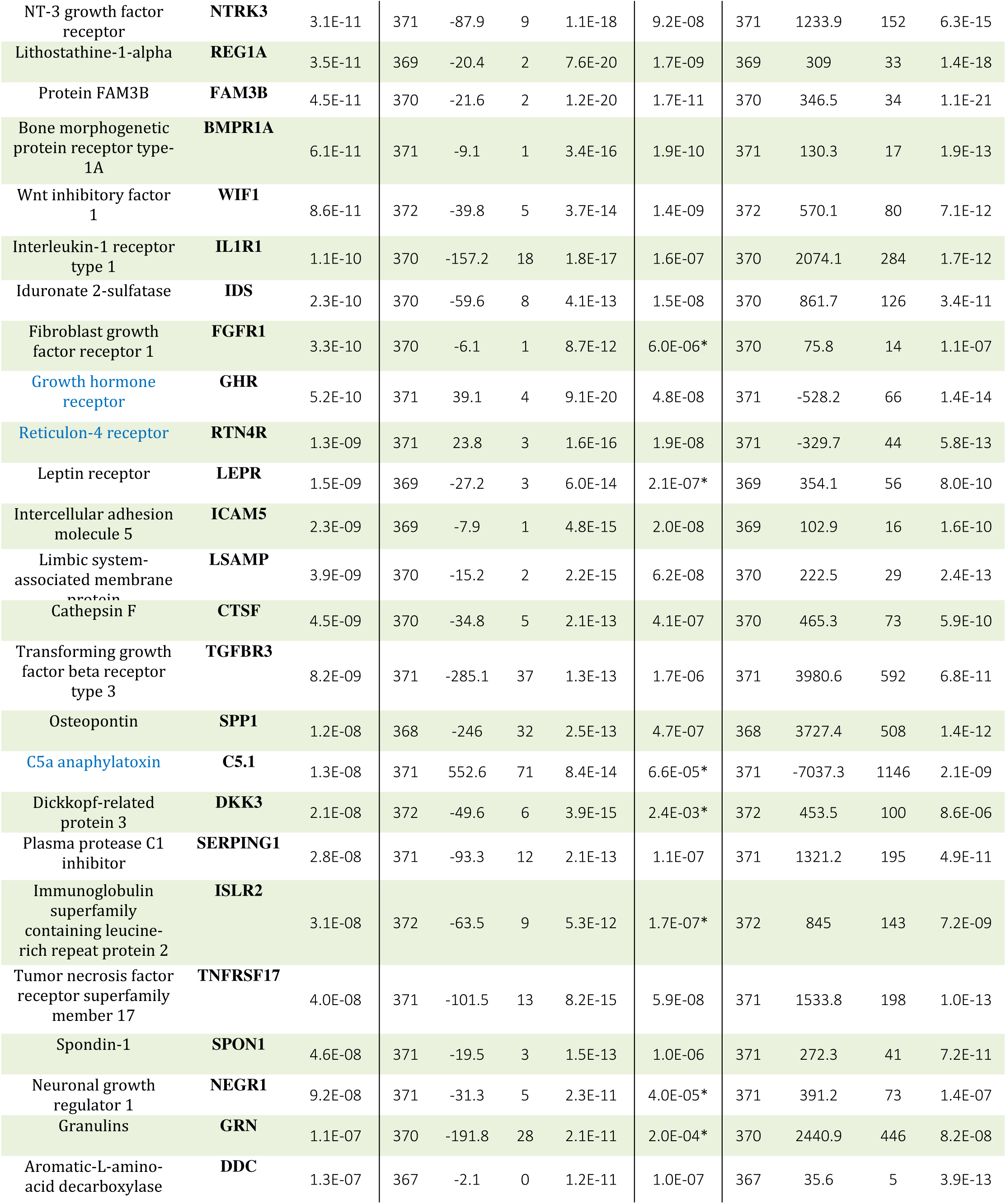
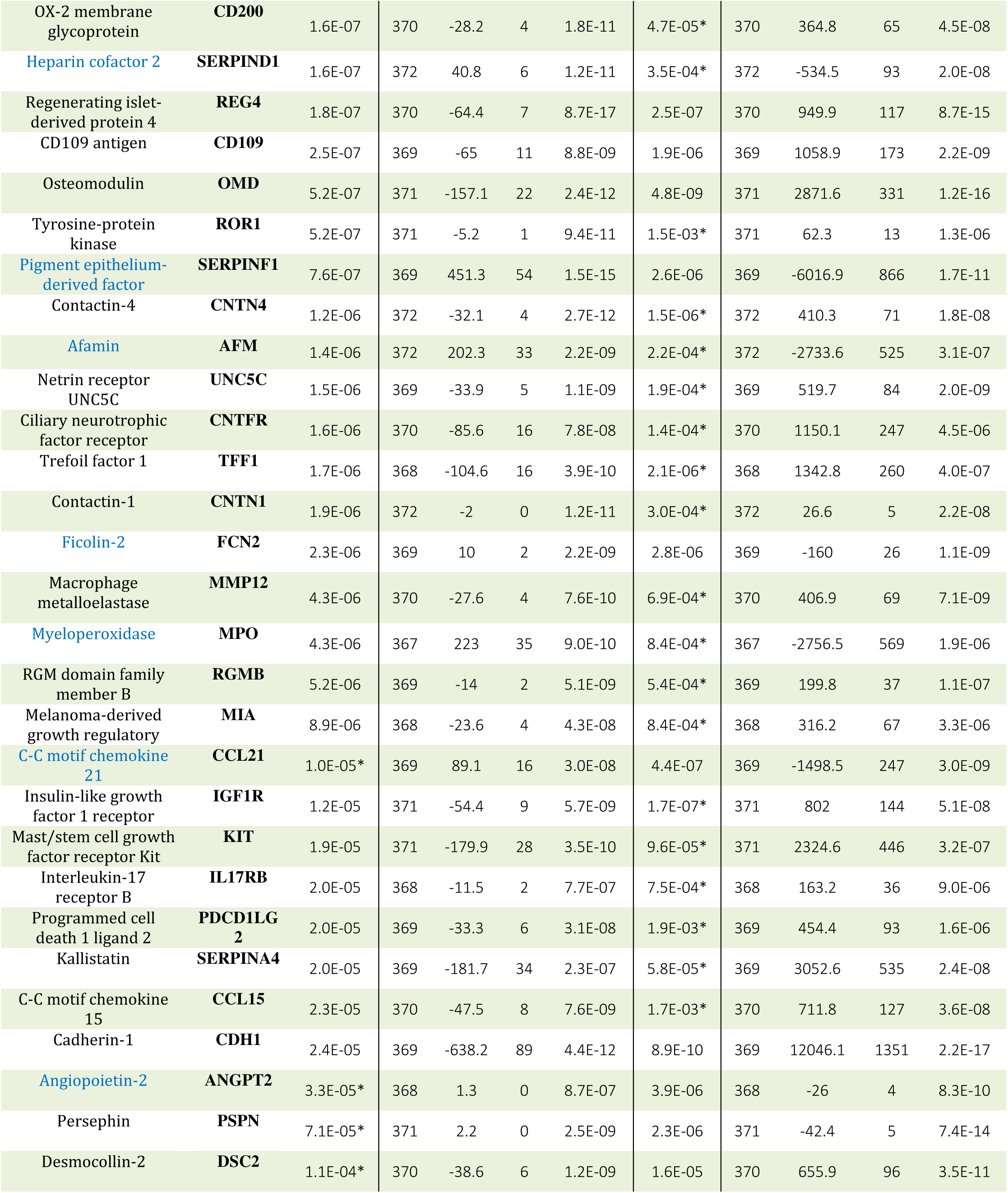
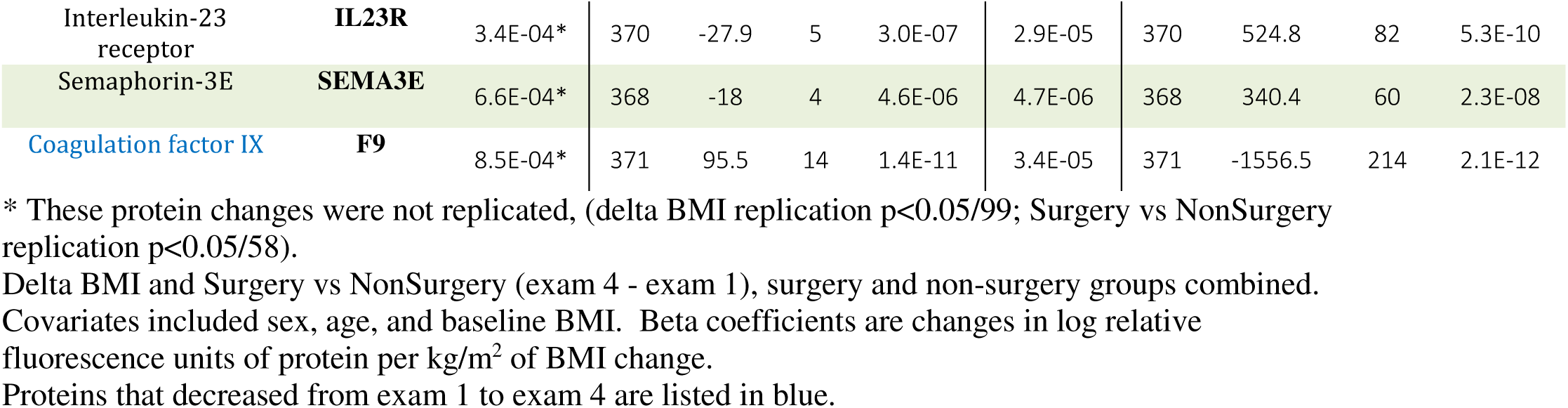
Regression results for protein changes versus BMI change and gastric bypass surgery versus non-surgery subjects (exam 4 – exam 1).

**Figure 1.**
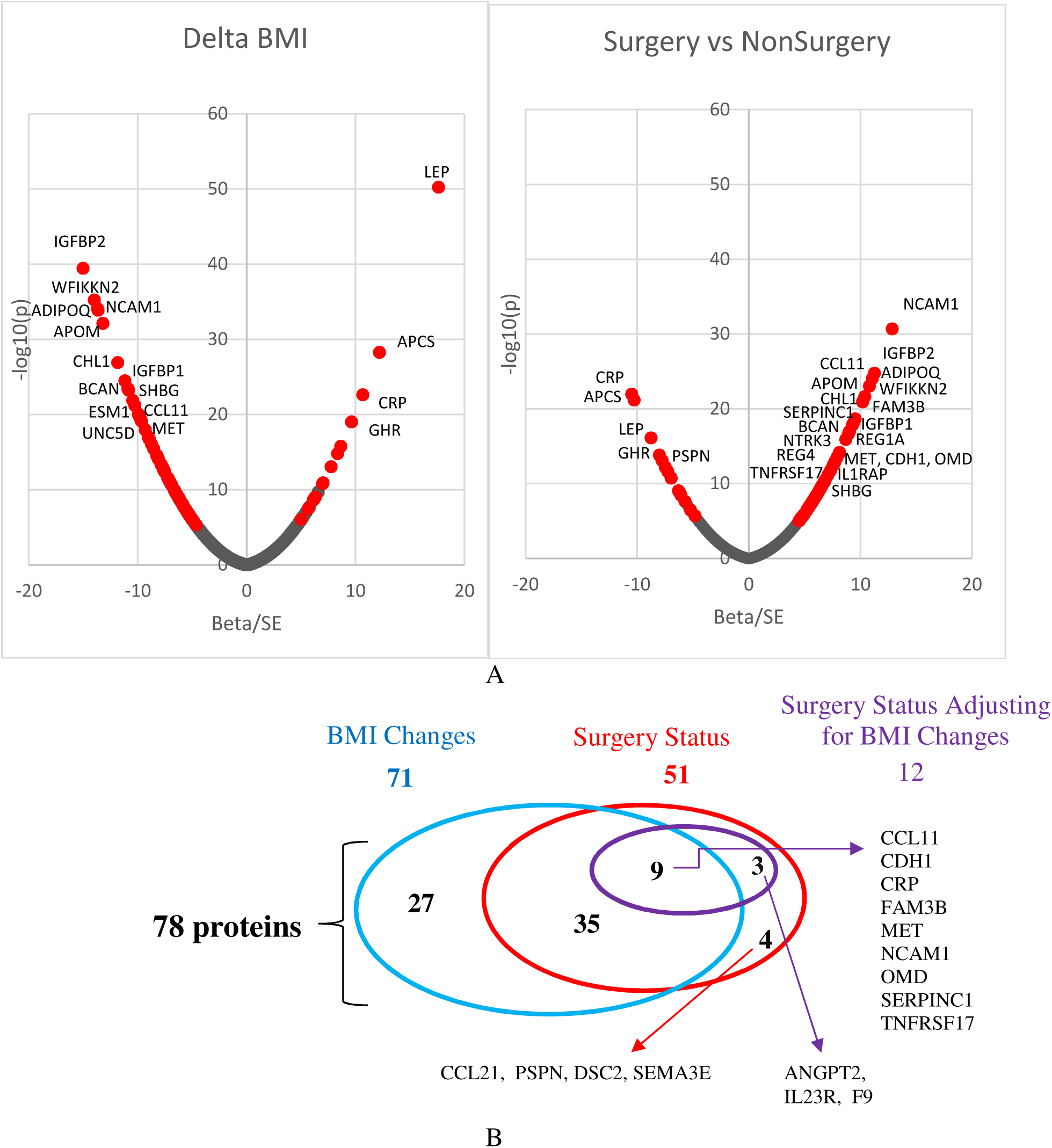
A) Plots of proteins’ normalized beta values (beta/SE beta) vs -log10(p values) from association results of delta BMI (exam 4 – exam 1) – left plot, and from surgery vs nonsurgery analysis – right plot, from combined discovery and replication samples. Red color is for the 78 significant protein changes. B). Overlap of 71 significant protein changes associated with BMI change (blue oval) and 51 associated with gastric bypass surgical status without correcting for change in BMI in the model (red oval). Twelve proteins of the 51 proteins were still significantly associated with surgery status after correcting for changes in BMI (purple oval).

### Seven proteins associate with differences between surgery and non-surgery groups but not with change in BMI

Twelve-year protein level changes in the gastric bypass surgery group were compared to the non-surgery group after adjusting for age, gender, and baseline BMI. A total of 58 protein changes were different between the two groups in the discovery set at a Bonferroni significance level (p<0.05/1297), among which 51 were replicated (p<0.05/58) (Table 2). Forty-four of those 51 proteins were also among the 71 proteins associated with change in BMI, while seven of these protein differences were not associated with change in BMI, including CCL21, ANGPT2, PSPN, DSC2, IL23R, SEMA3E, and F9 (Figure 1).

In addition, twelve of the 51 replicated protein changes remained significantly associated with surgery/non-surgery status after further adjusting for change in BMI (Figure 1). For example, change in CRP was significantly associated with change in BMI and the change differs between the surgical and non-surgical groups, but remained significantly different between groups even after adjusting for the change in BMI. Combining the significant results from the two analyses, 78 proteins changed significantly with either change in BMI or comparing surgery vs non-surgery between exams 1 and 4.

### Protein change correlations with BMI loss after 2 years and BMI regain from 2 to 12 years follow-up in the gastric bypass surgery group

Owing to this study’s longitudinal design where the same gastric bypass patient loses weight 1-2 years post-surgery but regains some weight during long-term follow-up, the consistency of the associations between protein changes and weight loss and weight regain during the two time periods could be compared. The 71 significant protein changes associated with changes in BMI (from Table 2) were subdivided into changes during the primary weight loss phase from surgery to 2 years post-surgery (exam 2) and during the weight maintenance or regain phase from exam 2 to exam 4 (12 years after surgery). Using only the surgery group (since protein levels were not measured at exam 2 in the non-surgical group), 38 of the 71 protein changes from Table 2 remained significantly associated with the 12-year changes in BMI at p<0.05/71 (Table 3). Of the 38 protein changes, 25 (65%) showed associations with BMI changes from exam 1 to exam 2 and 25 (65%) from exam 2 to exam 4, after Bonferroni correction for 38 × 2 tests (p<6.5 × 10^-4^). Twenty-one proteins were significant in both time periods. All 38 protein changes had the same direction of association with BMI change in both time periods, i.e., if the protein change was positively associated with BMI change from exam 1 to 2 (both decreased), it was positively associated from exam 2 to 4 (both increased) or if the protein change was inversely associated from exam 1 to 2 (BMI decreased but the protein increased), the protein change was inversely associated with BMI change from exam 2 to 4 (BMI increased but the protein decreased). Therefore, all 38 protein changes consistently tracked both BMI decreases and BMI increases.

**Table 3.** Association of changes in proteins with change in BMI in the gastric bypass surgery group

| Protein | Delta BMI: Exam 4- Exam 1 <sup>a</sup> |  |  |  | Delta BMI: Exam 2- Exam 1 <sup>b</sup> |  |  |  | Delta BMI: Exam 4 – Exam 2 <sup>b</sup> |  |  |  |
| --- | --- | --- | --- | --- | --- | --- | --- | --- | --- | --- | --- | --- |
|  | N | Beta<sup>c</sup> | SE | P-value | N | Beta<sup>c</sup> | SE | P-value | N | Beta<sup>c</sup> | SE | P-value |
| LEP | 192 | 1673.5 | 157 | 3.6E-21 | 191 | 1767.6 | 206 | 3.2E-15* | 190 | 1384.1 | 120 | 9.0E-24* |
| IGFBP2 | 191 | -43.2 | 5 | 1.5E-15 | 191 | -39.5 | 6 | 8.9E-11* | 191 | -51.3 | 5 | 1.1E-17* |
| ADIPOQ | 191 | -150.1 | 20 | 3.7E-12 | 191 | -182.6 | 23 | 5.9E-14* | 191 | -134.0 | 20 | 3.8E-10* |
| WFIKKN2 | 192 | -125.2 | 17 | 4.0E-12 | 191 | -84.9 | 20 | 3.5E-05* | 192 | -114.0 | 21 | 2.6E-07* |
| APOM | 192 | -311.3 | 46 | 1.4E-10 | 191 | -298.7 | 51 | 2.6E-08* | 191 | -177.6 | 47 | 2.3E-04* |
| GHRL | 190 | -21.2 | 3 | 4.7E-10 | 190 | -16.6 | 3 | 4.4E-07* | 191 | -16.8 | 4 | 7.8E-06* |
| TNFAIP6 | 189 | -16.6 | 3 | 4.9E-10 | 191 | -18.8 | 3 | 1.1E-08* | 190 | -17.3 | 4 | 1.8E-06* |
| APCS | 191 | 566.6 | 87 | 5.7E-10 | 192 | 752.6 | 133 | 5.3E-08* | 192 | 702.8 | 93 | 1.4E-12* |
| ESM1 | 191 | -24.9 | 4 | 8.0E-10 | 192 | -36.4 | 5 | 6.6E-11* | 191 | -21.5 | 4 | 5.3E-07* |
| SHBG | 191 | -173.0 | 28 | 5.1E-09 | 190 | -235.2 | 38 | 2.5E-09* | 190 | -171.5 | 29 | 1.4E-08* |
| DKK3 | 191 | -61.2 | 11 | 7.8E-08 | 192 | -75.8 | 14 | 2.4E-07* | 192 | -76.9 | 14 | 5.9E-08* |
| NCAM1 | 192 | -111.3 | 20 | 1.2E-07 | 191 | -81.7 | 27 | 2.7E-03 | 192 | -75.7 | 23 | 1.4E-03 |
| IGFBP1 | 191 | -51.3 | 10 | 2.1E-07 | 191 | -94.6 | 16 | 1.1E-08* | 191 | -49.7 | 14 | 3.5E-04* |
| CHL1 | 192 | -109.4 | 20 | 2.1E-07 | 192 | -148.4 | 28 | 2.4E-07* | 192 | -106.9 | 22 | 3.6E-06* |
| PIANP | 190 | -65.8 | 13 | 1.4E-06 | 192 | -60.5 | 17 | 4.5E-04* | 190 | -75.9 | 16 | 5.6E-06* |
| FGFR1 | 191 | -7.5 | 2 | 8.0E-06 | 191 | -10.5 | 2 | 1.2E-06* | 191 | -10.9 | 2 | 9.5E-09* |
| LSAMP | 191 | -15.3 | 3 | 2.0E-05 | 191 | -13.3 | 4 | 2.0E-03 | 191 | -19.4 | 4 | 1.5E-06* |
| ROR1 | 191 | -6.5 | 2 | 3.3E-05 | 192 | -6.7 | 2 | 3.7E-05* | 189 | -8.2 | 2 | 1.4E-07* |
| UNC5D | 190 | -75.0 | 18 | 3.6E-05 | 192 | -71.7 | 20 | 3.4E-04* | 189 | -106.1 | 18 | 3.2E-08* |
| NEGR1 | 191 | -37.2 | 9 | 5.3E-05 | 192 | -46.2 | 10 | 5.8E-06* | 191 | -45.3 | 9 | 1.6E-06* |
| CD200 | 190 | -30.4 | 7 | 5.4E-05 | 190 | -18.6 | 7 | 8.3E-03 | 190 | -30.2 | 8 | 8.4E-05* |
| GHR | 192 | 27.5 | 7 | 5.5E-05 | 192 | 30.1 | 9 | 1.2E-03 | 192 | 29.6 | 5 | 4.8E-08* |
| MET | 192 | -73.8 | 18 | 7.4E-05 | 191 | -94.8 | 23 | 4.3E-05* | 192 | -78.2 | 21 | 1.9E-04* |
| SERPIND1 | 192 | 41.7 | 10 | 7.7E-05 | 192 | 53.6 | 15 | 5.4E-04* | 192 | 29.2 | 9 | 1.5E-03 |
| CRP | 192 | 1321.5 | 332 | 9.6E-05 | 192 | 1004.9 | 474 | 3.5E-02 | 191 | 1443.7 | 325 | 1.5E-05* |
| NTRK3 | 192 | -67.7 | 17 | 1.1E-04 | 191 | -64.9 | 18 | 4.3E-04* | 191 | -61.9 | 18 | 5.7E-04* |
| C5.1 | 192 | 513.2 | 131 | 1.2E-04 | 191 | 426.7 | 172 | 1.4E-02 | 191 | 335.8 | 107 | 1.9E-03 |
| TGFB3 | 192 | -271.9 | 70 | 1.4E-04 | 191 | -304.5 | 79 | 1.6E-04* | 191 | -344.6 | 78 | 1.6E-05* |
| CCL11 | 191 | -154.1 | 40 | 1.8E-04 | 191 | -92.3 | 50 | 6.5E-02 | 191 | -64.3 | 46 | 1.6E-01 |
| CTSF | 192 | -36.6 | 10 | 2.0E-04 | 191 | -32.2 | 13 | 1.5E-02 | 192 | -33.3 | 12 | 4.8E-03 |
| MPO | 190 | 233.6 | 63 | 3.0E-04 | 189 | 206.8 | 70 | 3.3E-03 | 191 | 142.9 | 57 | 1.3E-02 |
| SERPINC1 | 192 | -625.5 | 170 | 3.1E-04 | 192 | -977.1 | 268 | 3.4E-04* | 192 | -416.8 | 201 | 4.0E-02 |
| BMPRI1A | 191 | -7.3 | 2 | 3.2E-04 | 191 | -6.7 | 2 | 1.3E-03 | 192 | -7.8 | 2 | 7.0E-04 |
| GRN | 190 | -179.6 | 49 | 3.3E-04 | 191 | -216.2 | 67 | 1.4E-03 | 190 | -145.1 | 58 | 1.4E-02 |
| **BCAN** | 190 | -18.6 | 5 | 3.6E-04 | 192 | -19.7 | 5 | 2.1E-04* | 190 | -17.0 | 6 | 4.9E-03 |
| **IL1RAP** | 191 | -273.5 | 77 | 4.8E-04 | 192 | -345.3 | 106 | 1.3E-03 | 191 | -155.7 | 76 | 4.3E-02 |
| **CNTN4** | 192 | -28.5 | 8 | 5.1E-04 | 191 | -56.6 | 11 | 2.6E-07* | 191 | -22.0 | 9 | 1.3E-02 |
| **SPON1** | 191 | -16.5 | 5 | 5.4E-04 | 191 | -18.8 | 5 | 7.5E-04 | 191 | -16.8 | 5 | 1.5E-03 |

\* Significant results for the changes from exam 1 to 2 or exam 2 to 4.
<sup>a</sup> After Bonferroni correction, p-values are significant if $p < 0.05/78 = 6.4 \text{ E-}04$ for exam 1 to 4 (38 proteins are significant).
<sup>b</sup> $p < 0.05/38/2 = 6.58 \text{ E-}04$ for time periods 1 to 2 and 2 to 4.
<sup>c</sup> Changes in proteins (log relative fluorescence units of protein change per $\text{kg/m}^2$ of BMI change) are exam 4 – exam 1, exam 2 – exam 1, and exam 4 – exam 2 respectively.

To visually compare the consistency of the amount of protein change with BMI change in the two time periods, the ratio of the beta-coefficients from the regression of protein changes on BMI changes from the two time periods was calculated (β_1-2_/ β_2-4_; Figure 2A). Ratios near 1.0 represent similar protein changes during weight loss as during weight regain, and strongly implicate weight change as the predominant factor for the protein change. Thirty-one of the 38 protein changes associated with BMI changes in the surgical group had a ratio within ±0.5 units away from the ideal value of 1.0 if their changes were similarly correlated with weight change in the two periods (Figure 2B). Seven proteins had a ratio > 1.5, suggesting that the per BMI unit changes in the protein during rapid weight loss from years 0 to 2 were greater than the changes from years 2 to 12. Those 7 proteins were APOM, ESM1, IGFBP1, IL1RAP, SERPINC1, SERPIND1 and CNTN4.

**Figure 2.**
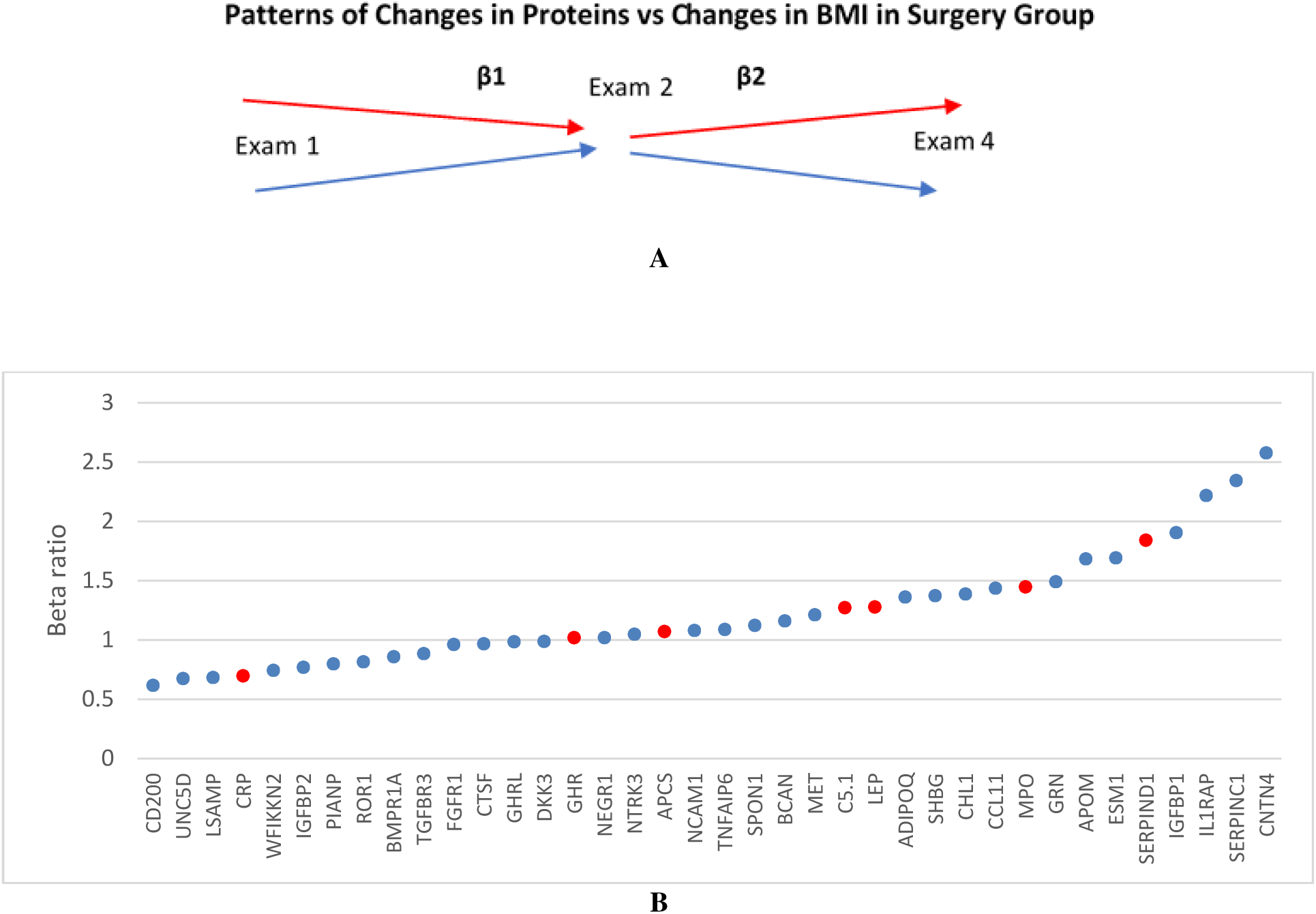
Patterns of protein change associations with BMI changes after weight loss (T2-T1) and after some weight regain (T4-T2). A) Follow up of the surgery group over two time periods shows 4 possible patterns: Pattern 1 (red): Proteins which decreased at T2 and then increased again at T4, i.e following the pattern of BMI change and producing a positive ratio of regression coefficients (P1-2/P2-4). Pattern 2 (blue): Proteins which increased at T2 and then decreased at T4, both time periods inversely related to change in BMI for a positive ratio. B) Ratios of two beta regression coefficients (P1-2/P2-4) obtained from surgery group (N=204 who have measurements at all 3 time points) for 38 proteins that replicated in surgery group from the total of 78 proteins. Marker colors correspond to the patterns in panel A.

### Proteins differentiating weight maintenance vs weight regain in the surgery group

Surgery subjects were divided into those who regained less than 10% of their baseline weight from exam 2 to exam 4 and subjects who regained more than 10% of their baseline weight (approximately the median weight regain and a previously suggested cutpoint for successful weight loss^16^). BMI at baseline, 2 years and 12 years was 45.1±6.6, 29.7±5.8, and 30.8±6.16 kg/m^2^ for those who maintained weight and 47.2±7.17, 29.6±5.5, and 39.03±8.5 kg/m^2^ for those who regained weight (Figure 3). There were significant mean differences between weight regain groups for 6 proteins (p< 0.05/1297; Supplementary Table 1). LEP and APCS were higher at exam 4 in those who regained more than 10% of their baseline weight while IGFBP2, WFIKKN2, HTRA2 and SHBG, were lower (Figure 3) and all 6 were among the 71 significant protein changes that correlated with BMI changes (Table 2).

**Figure 3.**
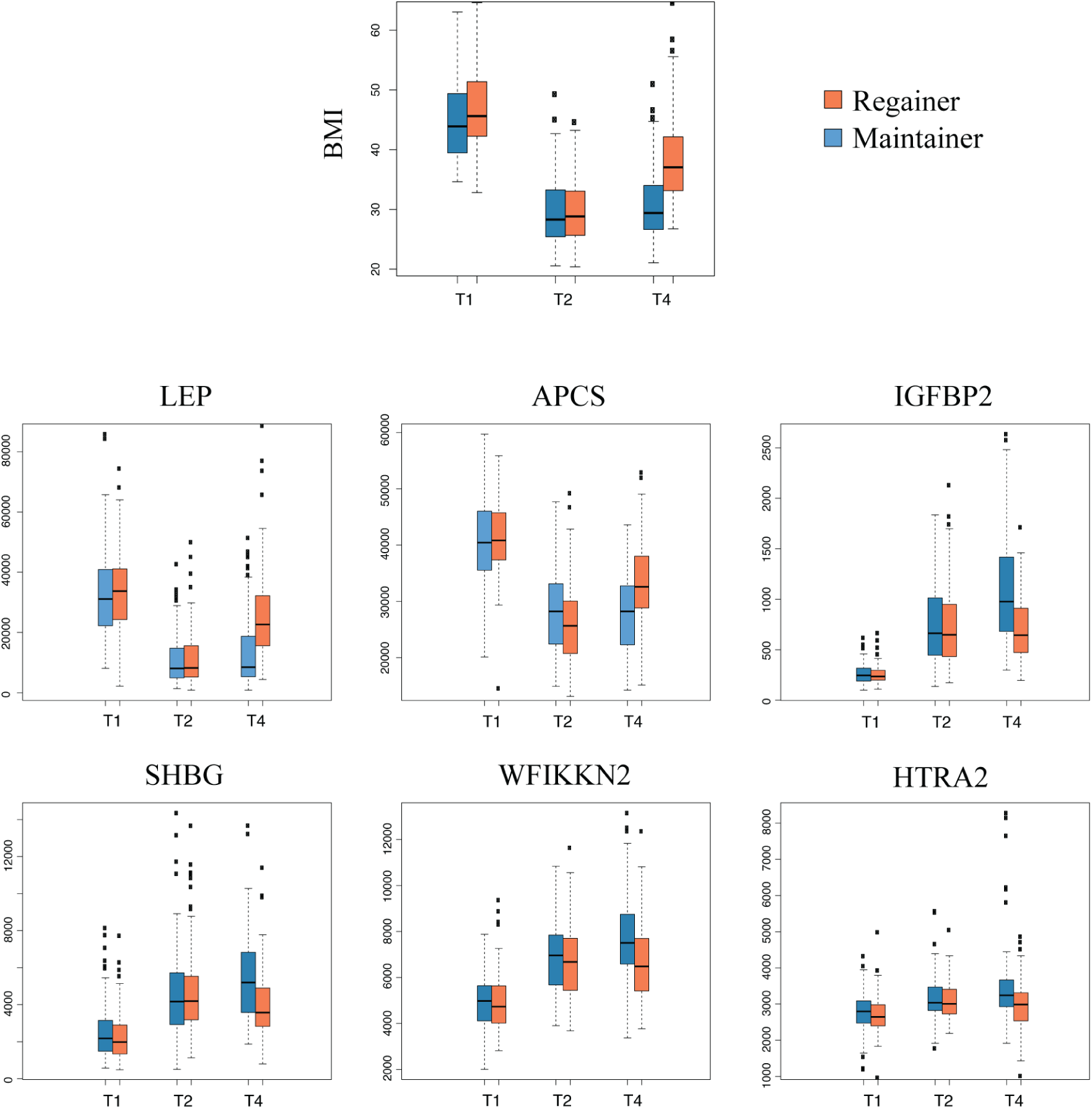
Boxplots at exams 1, 2 and 4 of BMI and the 6 proteins significantly different at exam 4 between subjects who maintained weight (blue, <10% of baseline weight regained from exam 2 to 4) and regained weight (red, >10% of baseline weight regained from exam 2 to 4).

### Clinical variable associations with protein changes

To investigate the clinical importance of the protein changes that were associated with BMI changes, 12-year changes in multiple clinical measurements (exam 4 – exam 1) were tested for association with the 71 proteins that were significantly associated with change in BMI (all subjects combined). The clinical variable changes were adjusted for age, gender, and BMI at baseline. A heat map of the regression beta-coefficients (sign(beta) × −1 × log10(p-value)) and the cluster patterns of the protein and clinical variable changes show BMI, FFM and FM form a cluster and have strong inverse associations with HDL-C (Figure 4). LDL-C, REE, uric acid and TG form a separate cluster. Glucose and HbA1c are more closely related to blood pressure and cardiovascular disease risk than the separate cluster of insulin, HOMA-IR, HOMA-B, DMINC and ALT. AST and DMREM. The set of 11 proteins that decrease with decreasing BMI clearly cluster together.

**Figure 4.**
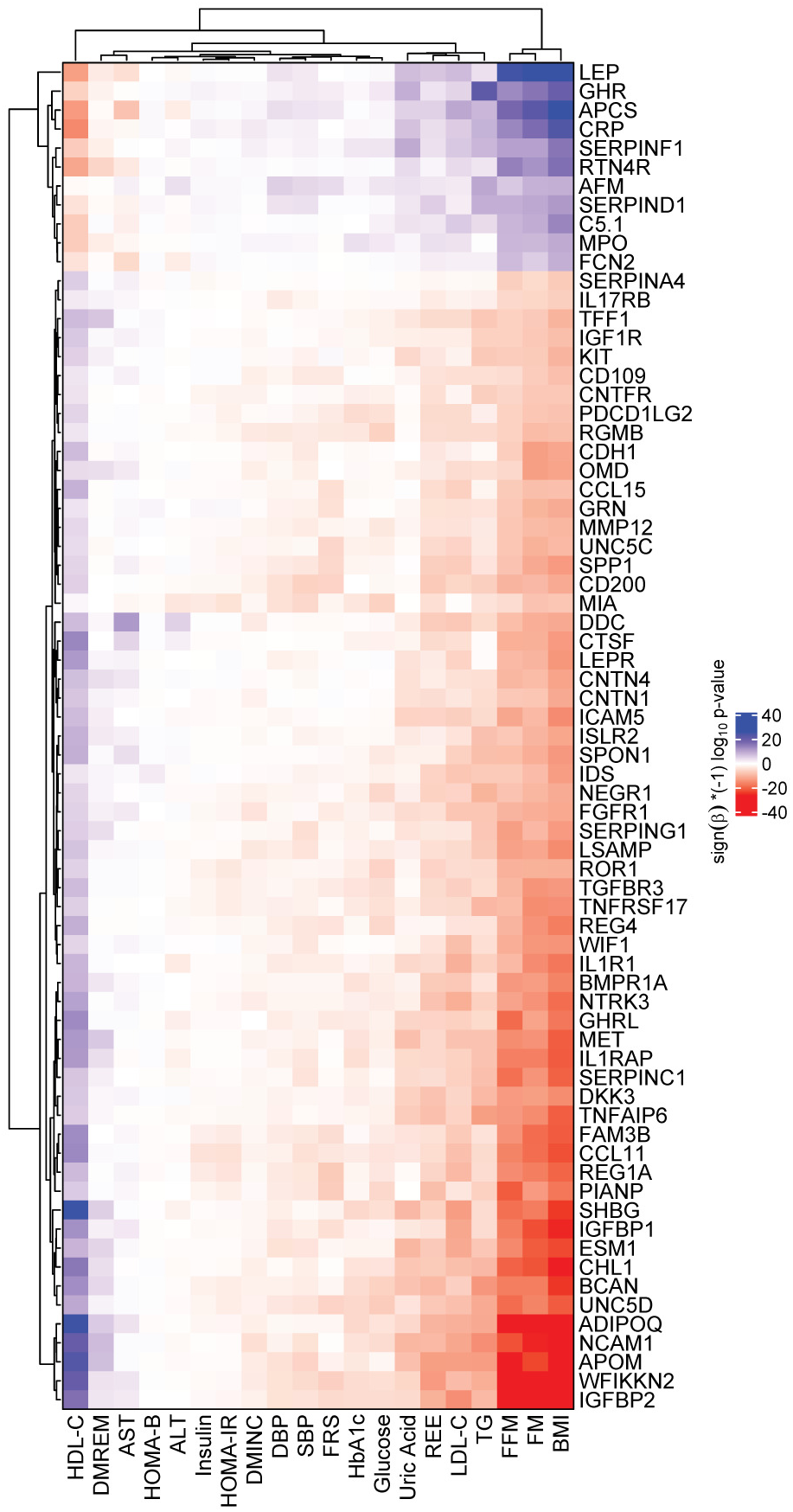
Heatmap of clinical variable associations with the 71 protein changes that were correlated with change in BMI for combined surgery and non-surgery groups. Values represent (sign(beta)*(−1)log10(p-value)) from a regression model. White indicates least significance, deepest red and deepest blue indicate highest significance with negative beta and positive beta, respectively. Clustering of the clinical variable and protein changes is shown. FM: fat mass; FFM: fat free mass; FRS: REE: resting energy expenditure; TG: triglycerides; SBP and DBP: systolic and diastolic blood pressure; Framingham risk score for 10-year CAD risk; DMINC and DMREM: diabetes incidence and remission.

The 71 protein changes related to BMI changes were most strongly associated with lipid-related proteins and not with variables in the glucose/insulin pathways. Seventy of the 71 protein changes were significantly related to FFM, all 71 to FM, 58 to HDL-C, 47 to LDL-C, 44 to TG, 38 to REE, 23 to uric acid, 8 to each of glucose, DMREM diabetes remission and FRS, while 6 or fewer were related to each of the remaining clinical variables (Figure 4). DMINC, HOMA-B, HOMA-IR and insulin had no significant associations with these proteins.

Because protein changes might have significant associations with non-lipid clinical variable changes independently of BMI changes (and therefore don’t appear in the list of 78 replicated proteins of Table 2), a heat map of associations of all 1297 protein changes with the clinical variable changes (exam 4 – exam 1) in the surgery group was created (Figure 5A). Despite including all proteins, longitudinal changes in the glucose, insulin, HOMA-B, HOMA-IR, HbA1c and blood pressure still do not show highly significant associations with protein changes. Analysis of the non-BMI change-related proteins showed significant associations with liver function, particularly proteins found in protein cluster 3 (Figure 5A).

**Figure 5A.**
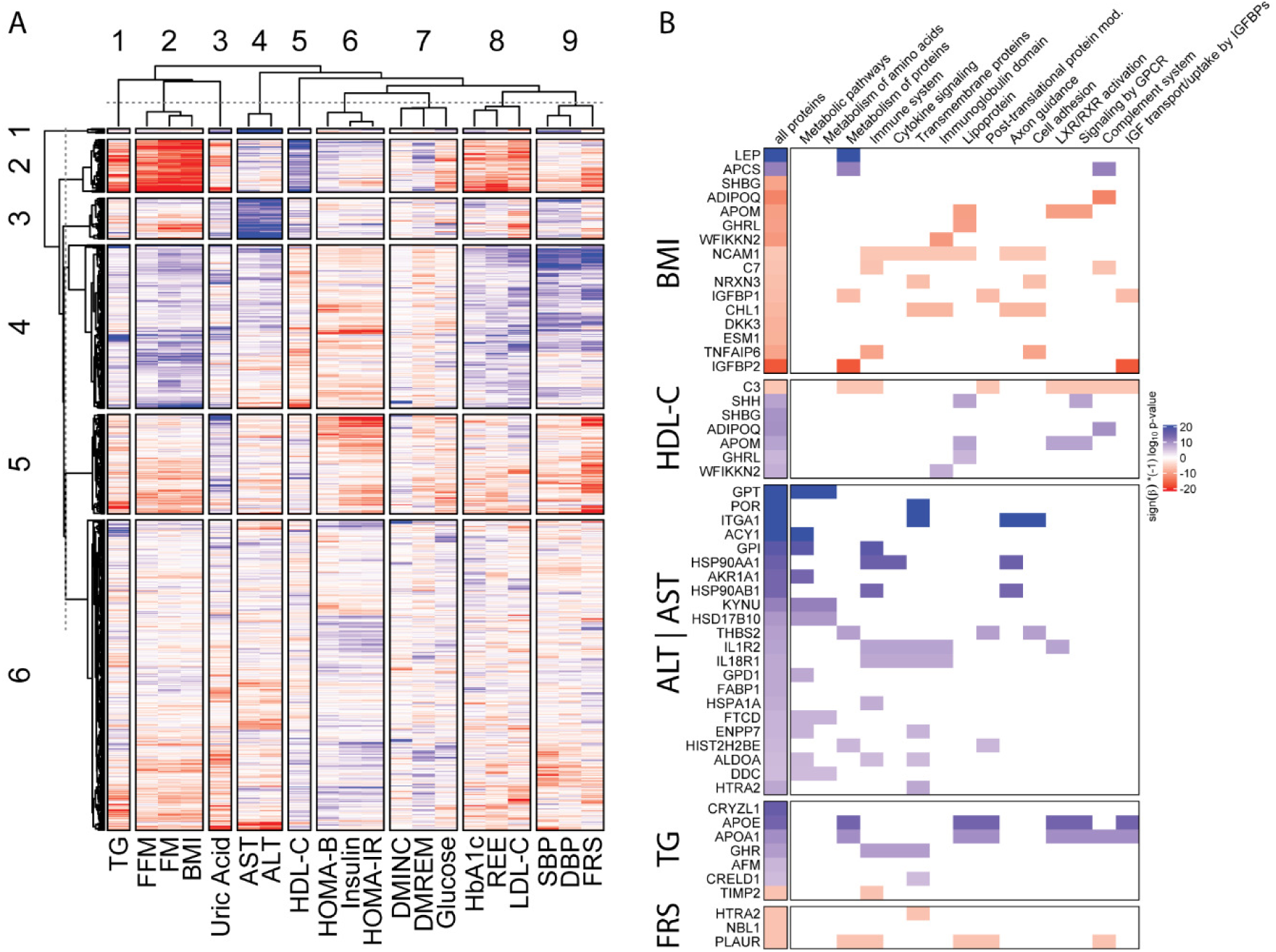
Heatmap showing significance of associations for all proteins with clinical variables. Red indicates high significance with negative beta (the protein increases with weight loss), blue indicates high significance with positive beta. Clustering of values results in 6 major clusters for proteins and 9 major clusters for clinical proteins. FM: fat mass; FFM: fat free mass; FRS: REE: resting energy expenditure; TG: triglycerides; SBP and DBP: systolic and diastolic blood pressure; Framingham risk score for 10-year CAD risk; DMINC and DMREM: diabetes incidence and remission. Figure 5B. Heatmap showing proteins with the most significant associations with a clinical variable and their functional annotation. Significance level is Bonferroni-corrected p-value (p = 0.05/1297/20) for 1297 proteins and 20 clinical variables. High (red) values indicate positive correlations with the clinical variables, meaning the protein level decreases with a decrease of the clinical variable. For example, the association of BMI change with leptin change is positive, indicating that leptin decreased with decreasing BMI. DMINC is not shown as it only has one protein (RBM39) significantly associated with it.

**Figure 6.**
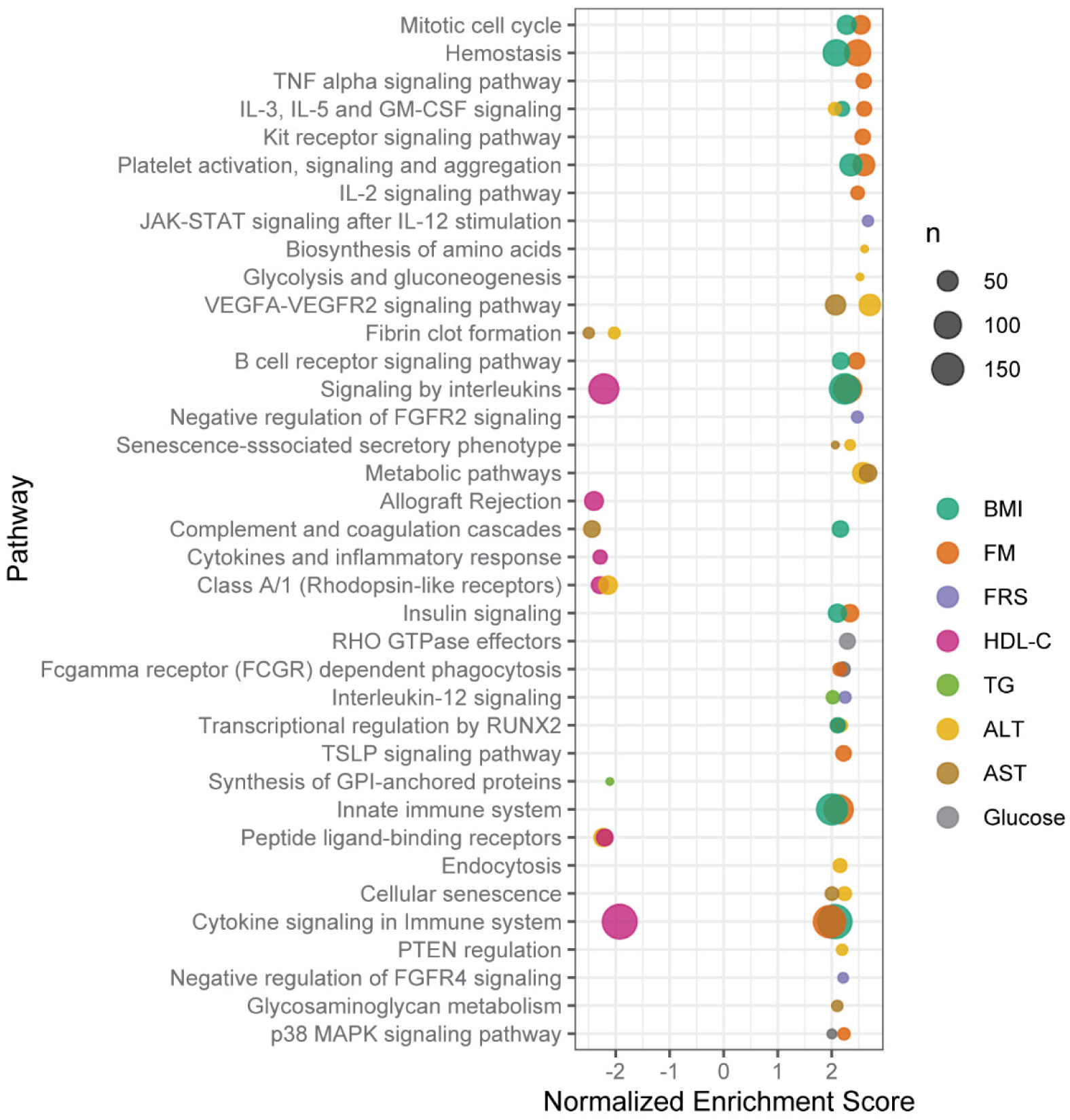
Dot plot showing enriched pathways within protein associations with clinical variables. Enrichment analysis was performed for the bypass surgery subset using GSEA. Normalized enrichment score (x-axis) indicates the distribution of pathway proteins within the distribution of sign(beta)*(−1*log10(p-value)) for the association of proteins with the clinical variable from a regression model as indicated by color. The size of this so-called core-enriched set is visualized by the size of the dots. Pathway enrichment cut-off for plotting is p < 0.022.

Figure 5B was derived from the data used in the heat map by selecting the 5 clinical variables that had one or more significant correlations with any of the 1297 protein changes at the more stringent Bonferroni adjusted level (-log10 of p < 0.05/(1297*20) = 5.71) and then showing the significance of those protein changes and the protein’s over-represented pathways. HDL-C change was the only clinical variable also significantly associated with any of the 16 proteins correlated with BMI change in Figure 5B, including ADIPOQ, APOM, WFIKKN2, GHRL and SHBG. SHH and C3 also were significantly associated with HDL-C changes, but not with BMI changes. There were 22 protein changes significantly associated with ALT/AST changes and these proteins were mainly in the metabolic pathways as would be expected. Lower TG is associated with lower AFM, GHR, APOA1, APOE, CRELD1, and CRYZL1, and higher TIMP2, none of which were associated with BMI change. There were three protein changes associated with the change in Framingham Risk Score (FRS) for ten-year risk of developing cardiovascular disease (CHD). These proteins were PLAUR, HTRA2 and NBL1, which all increased over 12 years and were inversely associated with risk change (decreased CHD risk). HTRA2, involved with apoptosis, is over-expressed in the transmembrane pathway while PLAUR2, a plasminogen activator protein, is over-expressed in multiple pathways including the complement and immune system pathways.

### Pathway analysis

Over-expressed pathways using gene-set enrichment analysis on the 78 proteins in Table 2 were identified using the SomaScan panel of proteins as background and are indicated in Table 4. Almost all pathways over-represented by the proteins involved inflammation, cellular growth, and apoptosis, systems which are altered with weight loss after bariatric surgery. These pathways were represented by the interleukins, MAPK signaling proteins, immune/complement system proteins, adipogenesis and insulin-like growth-related proteins. CRP, representing acute-phase inflammation, was limited to the complement pathway. In addition to identifying over-expressed pathways, there were 6 diseases that were over-represented by the significant protein changes (Supplementary Table 2). These diseases or conditions included bone mineral density, obesity/weight, metabolic syndrome and diabetes.

**Table 4.**
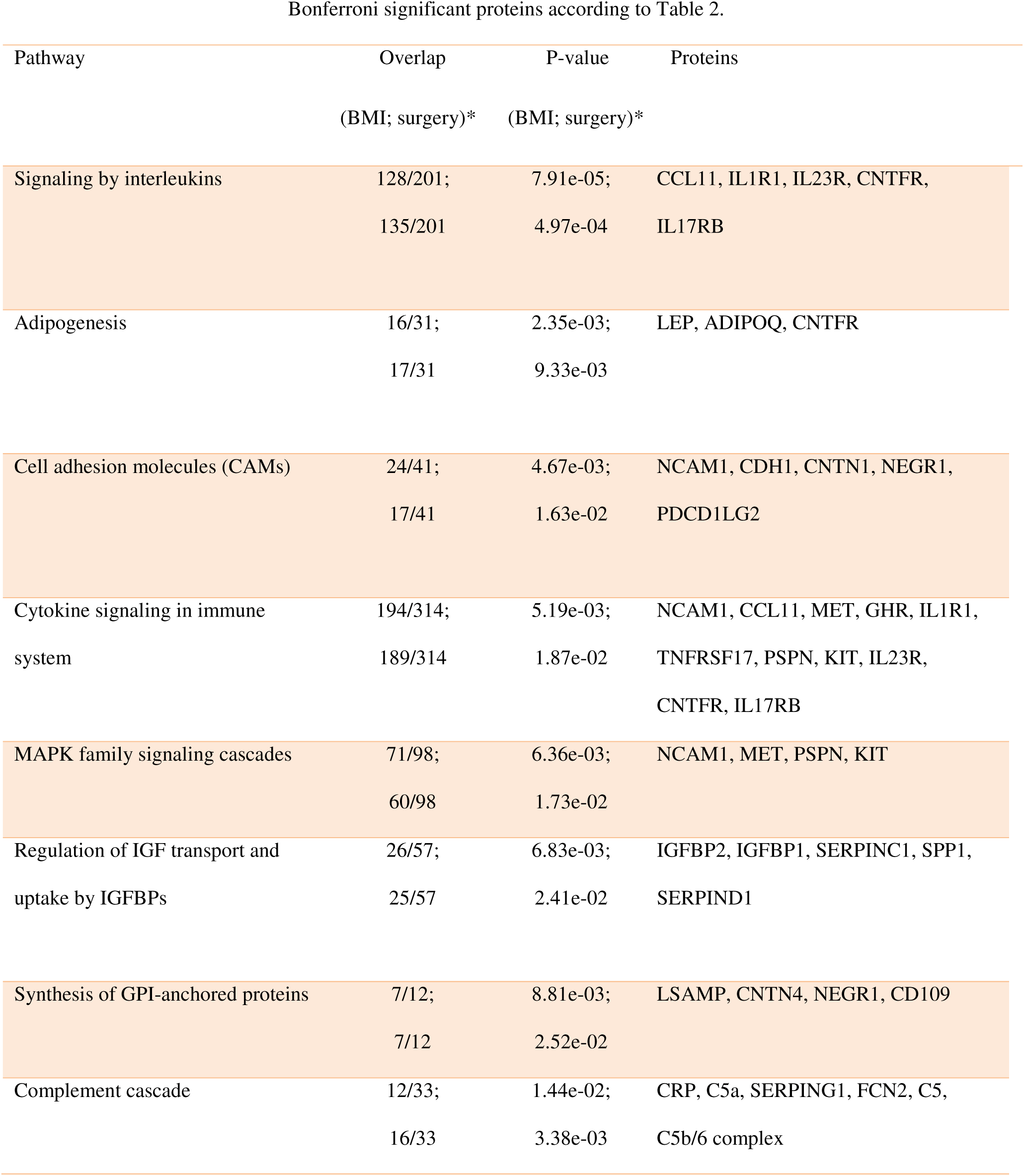
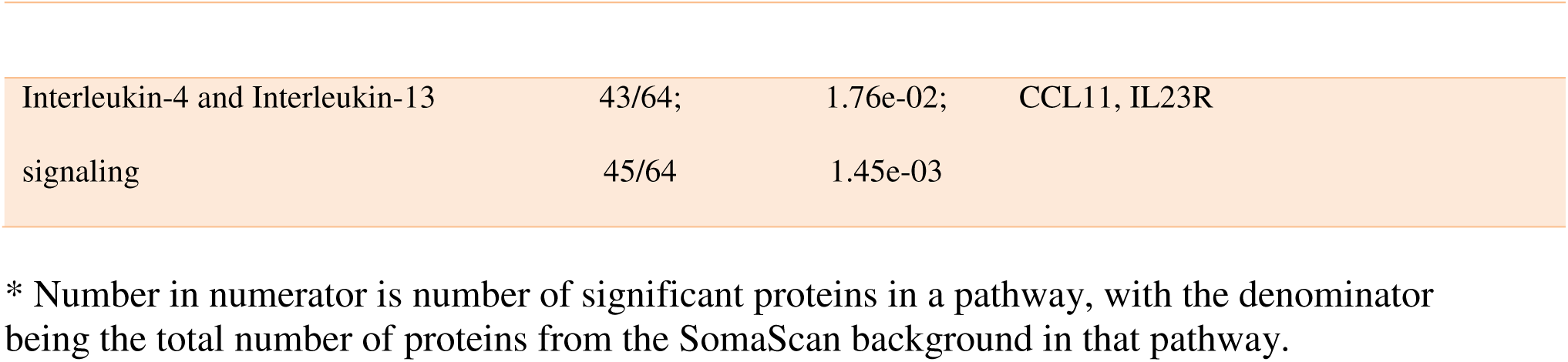
Top differential pathways identified by Gene-Set Enrichment Analysis (GSEA) of quantitative BMI change or surgery group status differences. Protein examples for each pathway are shown only if the protein was among the 78 Bonferroni significant proteins according to Table 2.

| Pathway | Overlap<br>(BMI; surgery)* | P-value<br>(BMI; surgery)* | Proteins |
| --- | --- | --- | --- |
| Signaling by interleukins | 128/201;<br>135/201 | 7.91e-05;<br>4.97e-04 | CCL11, IL1R1, IL23R, CNTFR,<br>IL17RB |
| Adipogenesis | 16/31;<br>17/31 | 2.35e-03;<br>9.33e-03 | LEP, ADIPOQ, CNTFR |
| Cell adhesion molecules (CAMs) | 24/41;<br>17/41 | 4.67e-03;<br>1.63e-02 | NCAM1, CDH1, CNTN1, NEGR1,<br>PDCD1LG2 |
| Cytokine signaling in immune system | 194/314;<br>189/314 | 5.19e-03;<br>1.87e-02 | NCAM1, CCL11, MET, GHR, IL1R1,<br>TNFRSF17, PSPN, KIT, IL23R,<br>CNTFR, IL17RB |
| MAPK family signaling cascades | 71/98;<br>60/98 | 6.36e-03;<br>1.73e-02 | NCAM1, MET, PSPN, KIT |
| Regulation of IGF transport and uptake by IGFBPs | 26/57;<br>25/57 | 6.83e-03;<br>2.41e-02 | IGFBP2, IGFBP1, SERPINC1, SPP1,<br>SERPIND1 |
| Synthesis of GPI-anchored proteins | 7/12;<br>7/12 | 8.81e-03;<br>2.52e-02 | LSAMP, CNTN4, NEGR1, CD109 |
| Complement cascade | 12/33;<br>16/33 | 1.44e-02;<br>3.38e-03 | CRP, C5a, SERPING1, FCN2, C5,<br>C5b/6 complex |
| Interleukin-4 and Interleukin-13 | 43/64; | 1.76e-02; | CCL11, IL23R |
| signaling | 45/64 | 1.45e-03 |  |

\* Number in numerator is number of significant proteins in a pathway, with the denominator being the total number of proteins from the SomaScan background in that pathway.

KEGG Brite pathway-based Voronoi treemaps, which require a protein to be assigned to only one pathway were also created for 950 out of the 1,297 panel proteins that could be assigned to a specific KEGG Brite level. Supplementary Figure 1 panels A, B and C, show absolute values of sign(beta) x -log10(p-value) for categories, pathways, and the proteins, respectively, associated with BMI changes. BMI changes were strongly associated with the Jak-STAT signaling pathway (panel B) in the signal transduction category (panel A). Changes in LEP, LEPR, GHR, and IL19 were the primary signal transduction pathway proteins. Change in BMI was also related to the complement and coagulation cascades, with C7, CFB and CFI being the primary proteins. The PPAR signaling pathway was represented by ADIPOQ. Pathways and proteins for TG, glucose, AST, ALT, FRS, and HDL-C are presented in Supplementary Figures 1B and 1C. In particular, the HDL-C beta values showed pathway and protein associations very similar to those seen for BMI (Supplementary Figures 1B and 1C).

## Discussion

As bariatric surgery is currently the only method that has achieved significant, consistent long-term weight loss, identifying the underlying reasons for risk factor improvements after weight loss has the possibility of developing better interventions for patients with severe obesity without the need for major surgery and bariatric surgery’s required life-long lifestyle changes. In addition, these interventions or treatments might be extended to those in the overweight or obese categories, especially for patients with significant co-morbidities such as diabetes. The large sample size and long follow-up period of this study allow us to test with sufficient power if the short-term protein changes observed in this and previous studies are maintained long-term, which maintenance is essential prior to pursuit of efforts to mimic the positive effects of these protein changes.

The major finding of this study of 378 individuals was that protein changes following gastric bypass surgery were durable 12 years post-surgery. The majority of the protein changes occurred within 2 years of surgery and tended to reverse after the first two years according to the amount of weight regain during the following 10 years. These are important results as they support the clinical data that the health benefits of gastric bypass surgery occur soon after the surgery rather than following a significant period of time. The protein changes during the weight regain period remained small, despite the statistical significance of the changes, as there were only six proteins that significantly differed at 12 years between those who regained less than or more than 10% of their initial weight. Larger protein changes during weight loss compared to weight regain periods were seen for APOM, ESM1, IGFBP1, IL1RAP, SERPINC1, SERPIND1 and CNTN4, possibly suggesting non-linear associations with BMI change or additional BMI-independent mechanisms that are activated during rapid weight loss.

Another important finding of the study is that most of the changes in proteins were associated with changes in BMI per se and not BMI-independent pathways induced by the surgical rerouting of the intestine. Only 16 of the 78 replicated protein changes remained significant between the surgical and non-surgical groups after adjustment for changes in BMI. The ratio of the regression beta-coefficients of protein change with BMI change suggested most proteins changed approximately the same per unit BMI change regardless of whether weight was lost or weight was regained. These paired within-person results strongly supported the involvement of BMI change with these protein changes. Well-known proteins previously implicated with obesity were confirmed to also be related to changes in obesity during both weight loss and weight regain, including those shown in Table 5B such as LEP, SHBG, ADIPOQ, GHRL, IGFBP1, and IGFBP2. WFIKKN2 was associated with BMI, and over-expression in mice has been associated with developing increased muscle mass^17^. The increased expression seen during weight loss may occur to counteract the lean mass lost after gastric bypass surgery.

The majority of protein changes that were significantly associated with BMI changes were related to changes in lipid levels as opposed to blood pressure, liver function, or glucose- and insulin-associated variables. After expanding the analyses to all protein changes regardless of their correlation with BMI, diabetes-related clinical variable associations with protein changes remained conspicuously absent, and those that did appear were not the same proteins related to BMI changes. Diabetes, hypertension, and NAFLD significantly decrease following gastric bypass surgery even after 10-12 years^4,18^. However, these diseases did not seem to be related to the protein changes associated with BMI change or surgery status and only minimally associated with any of the 1297 protein changes. The lack of strong associations of diabetes-related variables with obesity-related protein changes could arise if the relevant proteins were not included in the panel of proteins tested, but these results are otherwise unexplained.

### Specific protein decreases after gastric bypass surgery

Of the 78 proteins identified, 11 proteins decreased with the 12-year decrease in BMI in accordance with some of their known functions. Decreasing LEP is highly correlated with weight loss^19^. Weight loss is known to reduce inflammation in accordance with the reduction in CRP, C5.1, and MPO seen in our study. As tissue growth is being opposed by the greatly reduced diet and intestinal reabsorption of nutrients, there is less need for GHR, which decreased. AFM (afamin) was decreased with weight loss, confirming the higher levels of AFM being associated with increased weight, the metabolic syndrome^20^ and DM incidence^21^. APCS is thought to bind to and remove apoptotic cells. At 2 years when the large tissue losses during the first 2 years have stabilized, there may be less need for damaged cellular removal, as APCS is strongly decreased at 2 years after surgery. One of the functions of RTN4R is to protect against apoptosis, which seems to counteract APCS. However, this RTN4R action may act only on neuronal tissue such as motoneurons and its relationship to actin cytoskeleton reorganization may be the more important function. The actin filament system was the most over-expressed pathway during weight loss and gain in the study of Piening, et al^22^.

FCN2 is involved with innate immunity and lectin complement pathway. The complement system has been previously shown to be altered after gastric bypass surgery^11,23^. SERPINF1 is released by adipocytes^24^ and with the decrease in adipocytes after surgery, SERPINF1 showed significant decreases at 12 years. It inhibits angiogenesis, the need for which would be reduced with a reduction in tissue area. It is not clear why SERPIND1 is reduced after gastric bypass surgery as the *SERPIND1* gene codes for heparin cofactor II, is a thrombin inhibitor and has been suggested to protect against atherosclerosis^25^. However, higher SERPINC1 was highly inversely correlated with BMI. The *SERPINC1* gene codes for antithrombin-III and when antithrombin-III is elevated its presence may become dominant over heparin cofactor II (https://www.uniprot.org/uniprot/P05546#function). Both SERPINC1 and SERPIND1 were both down regulated in NAFLD^26^ whereas in this study SERPIND1 was down regulated with weight loss, suggesting the SERPIND1 signal may not be very robust while SERPINC1 increased as expected.

### Specific protein increases after gastric bypass surgery

Some of the proteins with the most significant increases with decreasing BMI were IGFBP1 and IGFBP2, ADIPOQ, and APOM. IGFBP1 and IGFBP2 are insulin-like growth factor binding proteins and higher levels have been associated with reduced glucose intolerance, blood pressure or NAFLD in multiple cross-sectional or short-term intervention studies^11,26-29^. Therefore, the increases seen 12-years after surgery may be partially responsible for, although they were not significantly related to, the remission of diabetes in the surgical patients. ADIPOQ has multiple beneficial functions including improvement in most components of the metabolic syndrome, reducing diabetes and atherosclerosis^30^. APOM is a component of HDL and is thought to be responsible for the anti-inflammatory effects of HDL. The higher levels of APOM after bariatric surgery correspond to the reduced inflammation present after weight loss^31^, which is also supported by a significant association of ApoM with remission of diabetes. Since denser HDL particles are the primary carriers of APOM, one might also expect a shift to more dense HDL particles with increasing APOM after surgery. However, the significant increase in cholesterol content in the HDL particle along with lower ApoA1 at follow-up (Surgery group: Exam 4 – Exam 1: T statistics: −6.33, p-value: 1.53E-09, and Exam 2 – Exam 1, T statistics: −4.46, p-value: 1.32E-05) suggests that the particles were less dense. ApoE and ApoB, though not associated with weight loss, decreased from baseline to 12 years in the surgery group as would be expected with lower triglycerides and LDL-C (non-significant) and confirms weight loss associations with multiple apolipoproteins seen in another weight loss study^32^.

### Clinical variable associations with the 71 replicated proteins and all 1,297 proteins

HDL-C was clearly the mostly strongly associated clinical variable associated with the 71 protein changes that were associated with BMI change after surgery. HDL-C also was alone in its own cluster of the clinical variables (Figure 4). HDL-C increased by approximately 25% two years after surgery, which increase was maintained throughout the 12-year follow-up period despite weight regain and was larger than any other clinical change^4^. HDL-C has a strong positive association with SHBG even after adjusting for multiple cardiovascular risk factors^33^. Polymorphisms in the *SHBG* gene have been associated with abundance of the SHBG protein, HDL-C, diabetes, and CHD^33-35^. Additionally, decreases in AFM, APOE (associated with decreased TG), and increases in ADIPOQ and APOM (associated with increased HDL-C) after surgery suggest these proteins may be responsible for the sustained large increase in HDL-C. These associations also suggest that CAD risk should be lowered^36^ 12 years after surgery. However, none of these protein changes were associated with the 10-year risk of future CAD assessed by the Framingham Risk Score. Most protein associations with the clinical variables seemed to be highly specific (little pleiotropy) for only one clinical variable. For example, protein associations with TG were confined to TG and not significantly related to the other clinical variables (Figure 5B).

### Pathways

The gene-set and Voronoi treemap analyses used for pathway analysis allowed us to identify several pathways associated with weight loss and the inflammation due to obesity. Major pathways associated with BMI are related to lipid metabolism, adipose tissue functionality, inflammation, cell adhesion, immunity and coagulation. Reduced inflammation appeared to be a primary benefit of weight loss and gastric bypass surgery, as suggested by the over-expression of protein changes in the pathways involving signaling by interleukins, cytokine signaling in the immune system, and more specific interleukin pathways. These pathways coincided with the over-expressed HDL-C pathways supporting the anti-inflammatory functions of elevated HDL-C. The cytokine signaling pathway contains a number of interleukins and adipocytokines, and adipose tissue colonization by pro-inflammatory macrophages and macrophage-derived factors like TNF-α and IL-β is a hallmark of obesity^37,38^.

Three important pathways were identified using Voronoi treemaps. One pathway was the AMPK pathway, which regulates cellular energy stores and preservation of ATP and is activated by leptin and adiponectin^39,40^. One would also expect this pathway to be activated after the substantially reduced dietary intake required after surgery^41^. It can inhibit gluconeogenesis and lipid and protein synthesis. Another pathway, JAK-STAT, can be activated by cytokines^42^ and leptin, and functions include regulating cellular division and inflammation processes and proper adipose function. A third pathway, PIK3-AKT-ras pathway, influences cell survival and growth, is involved with lipid phosphorylation and has been associated with diabetes^43^. Additional pathways involve cell adhesion molecules and complement and coagulation cascades that affect coagulation and fibrin formation^44^. Many of those pathways also underlie control of the clinical variables used in this study (AST, ALT, TG, HDL-C and FRS). Amino acid metabolism and glycolysis are more pronounced for AST and ALT, as expected, due to the greatly altered liver metabolism after bariatric surgery.

These pathway results and the associated proteins support the idea that the dramatic improvements in health after bariatric surgery arise from improvements in multiple systems and proteins. The results replicate those found in a short-term weight gain and weight loss study^22^. Coagulation, complement, and inflammation proteins were in primary pathways involved with weight gain and were reversed with weight loss, similar to our long-term study. Findings in a study of 47 subjects (19 of whom were followed at least 2 years) who had gastric bypass surgery were also similar, showing apolipoproteins in particular (including APOM) with the most significant changes, along with complement and inflammation pathways^11^.

## Strengths and Weaknesses

One of the known weaknesses of proteomic platforms such as SomaScan and OLINK in measuring protein abundance is that it has been documented that DNA polymorphisms in the region of the aptamer binding can affect the assay^45-47^. No attempt was made to investigate this in this study, but most of the comparisons used were paired measurements of changes in a person over time. Therefore, if deficient binding occurred, it did so for both the baseline and follow-up samples. It is not clear how these polymorphisms might affect the results of such paired testing in this study.

While there was an internal replication protocol and most proteins in the discovery set were replicated, the results would be even more convincing with an external replication cohort. Such studies with 12-year follow-up or even 5-year follow-up have not been reported for proteomics. Nevertheless, there was an overlap of many of the proteins identified in this study with the very low-calorie diet intervention study of Geyer, et al^32^, the weight gain and loss study of Piening, et al^22^ and the gastric bypass study of Wewer Albrechtsen, et al^11^.

Strengths of the study include the large sample size, long follow-up, and having a non-intervened, severely obese comparison group to account for changes due to aging, changes in diet or exercise, or onset of disease.

## Conclusions

Significant short-term changes in proteins seen up to 2 years after gastric bypass surgery remained significantly changed 12 years after surgery despite some weight regain occurring. Most protein changes found associated with BMI changes in the discovery sample were replicated in a second sample and the reversal of direction of protein changes after weight loss when weight was partially regained suggest our findings were robust.

Most protein changes were related to changes in BMI rather than non-weight related long-term surgical effects suggesting that the beneficial changes of these proteins can be obtained by weight loss rather than bariatric surgery. These BMI-associated protein changes were associated with lipid-related proteins and pathways that control inflammation, energy conservation, cell replication and apoptosis, and adipocyte function. The non-BMI related protein changes were related to inflammation, coagulation and organismal injury pathways.

The clinical variables most associated with the replicated proteins included HDL-C and to some extent other lipids. The direction of all of these protein and clinical variable changes would be expected to produce the health benefits and decreased mortality observed after bariatric surgery. Mechanisms responsible for sustained higher HDL-C (and lower ApoA1) after gastric bypass surgery, particularly their anti-inflammatory functions, and the related protein increases of ApoM, SHH and GHRL in the lipoprotein pathway and decreases in C3 in multiple pathways need to be clarified. The expected associations of weight changes with LEP and ADIPOQ were strongly replicated as were other proteins previously related to obesity, such as SHBG, IGFBP1, IGFBP2 and AFM. Other proteins not previously suggested by cross-sectional or short-term longitudinal studies were found to be strongly related to BMI change after 12 years and need further replication and investigation.

## Methods

### Materials

Starting in 2000, a longitudinal study of the risks and benefits of Roux-en-Y gastric bypass surgery was begun. After a baseline exam (exam 1), additional exams were conducted at approximately 2 years (exam 2), 6 years (exam 3) and 12 years (exam 4). Full details have been previously published^4,48,49^. There were 1156 subjects enrolled in the study, of whom 418 had gastric bypass surgery and 738 did not have bariatric surgery within one year of the baseline exam. A subset of the subjects who had gastric bypass surgery were identified who had a blood draw at baseline and at exam 4, 12 years after surgery. Subjects who never had bariatric surgery during the 12 years of follow-up and had baseline and 12-year follow-up blood draws were also selected. The initial set of subjects used for plasma protein measurements was 203 subjects (137 surgery and 68 non-surgery). Because of additional funding, a second subset of subjects was later selected and used to replicate the findings from the first subset of subjects and comprised 175 subjects (97 surgery and 78 non-surgery). Plasma proteins were also measured for 204 of the 234 subjects who had gastric bypass surgery who also had an available blood sample from exam 2. Proteins from exam 3 were not measured on any subject. All subjects provided written informed consent and the study was approved by the University of Utah Institutional Review Board.

### Clinical Measurements

Weight was measured by a Scaletronix scale (model 5100; Scaletronix Corp., Wheaton, IL). The scale has an accuracy of 0.1 kg with a limit of 364 kg. A Harpenden anthropometer (Holtain, Ltd., Crymych, UK) was used to measure height to the nearest centimeter. Change in BMI was calculated as follow-up BMI minus baseline BMI. Fat free mass (FFM) and fat mass (FM), measured by bioimpedance, glucose, insulin, HOMA-IR, high density lipoprotein cholesterol (HDL-C), low density lipoprotein cholesterol (LDL-C), triglycerides (TG), aspartate aminotransferase (AST), alanine aminotransferase (ALT), resting metabolic rate (RMR), systolic and diastolic blood pressure (SBP, DBP), diabetes remission at 12 years in baseline diabetics (DMREM), diabetes incidence in baseline non-diabetics (DMINC) and 10-year risk of coronary heart disease (CHD) by the Framingham Risk Score (FRS) were measured or calculated as previously described^4,48-50^. So that the Framingham Risk score represented change in CHD risk only due to changes in the clinical variables and not age, age at exam 4 was used in both the baseline and 12-year follow-up risk equations. Diabetes was defined as a fasting glucose of 126 mg/dl or greater or being on diabetic medication.

Blood was drawn at each exam, separated, and plasma aliquots were stored at −80 degrees C. Prior to assay, the plasma was thawed and approximately 70 ^l used for the proteomics assay. Proteins were measured by a Slow Off-rate Modified Aptamer-based protein array (SomaLogic, Boulder, Colorado)^51^. A total of 1,297 proteins were measured and are referred to by their gene names. Normalization and calibration of protein levels were done using SomaScan proprietary software. Sample data was first normalized to remove hybridization variation within a run, followed by median normalization across all samples to remove other assay biases within the run and finally calibrated to remove assay differences between runs. Calibrator CVs on each plate were calculated and at least 50% of SOMAmer reagents had to have CVs less than 0.1 and 95% had to have CVs below 0.2. Any flagged samples by SomaScan software were removed from the analysis, resulting in 203 subjects in the discovery batch and 175 subjects in the replication batch, for a total of 378 study subjects. Protein abundance was calculated as log of the relative fluorescence units.

### Statistical methods

Within individual changes in protein levels (protein at exam 4 – protein at exam 1) were calculated for all 1,297 proteins. Four major hypotheses were targeted: Regression models were used to test for protein changes versus BMI changes (exam 4 – exam 1) or versus surgery status (surgery group compared with non-surgery group), after adjusting for age, gender, and baseline BMI, first in the discovery batch and second in the replication batch; Clinical variable tests were done using only the gastric bypass surgery group in the analysis with both discovery and replication subsets combined. Ratios of the regression beta coefficients from the two time periods were created using the associations of BMI change for protein changes from exam 1 to exam 2 versus protein changes from exam 2 to exam 4: ratio = beta (exam 2 - exam 1) / beta (exam 4 - exam 2). These ratios represent whether or not the association of change in BMI had the same, smaller, or larger effect sizes for the weight-loss time period versus the weight-regain time period.

Outliers were removed at 4 standard deviations above or below the mean value of each protein. All statistical tests were two-sided tests. For hypotheses 1-2, protein change correlations were considered significant at Bonferroni-adjusted p-value of p<0.05/1,297. The replication p-value threshold was p<0.05/99 as there were 99 significant proteins found in the discovery phase. For hypothesis 3, the adjusted p-value for significance was considered to be p<0.05/51, for the 51 protein changes found significant. For hypothesis 4, there were 78 proteins tested only in the surgical group (p<0.05/78), and of the resulting 38 that were significant for exam period 1 to 4, there were 2 additional tests for periods 1 to 2 and 2 to 4 (p<0.05/38/2).

Clinical data was winsorized at 5-95% quantiles and tested using linear models. A secondary analysis of the clinical variables was performed using all 1,297 proteins (rather than just 78 replicated proteins). P-values were considered significant for the 20 clinical variables when p<0.05/1,297/20.

To help identify biological pathways involved with the beneficial health changes seen after gastric bypass surgery, the 78 significant and replicated protein changes were tested for association with other clinical variables, including metabolic, blood pressure, and diabetes phenotypes in the combined surgery and non-surgery subjects. Linear regression models were used as above with covariates age, gender, baseline BMI, and a batch term. Blood pressure, lipids, and diabetes-related variables that are affected by antihypertensive, antidiabetic or lipid medications were adjusted prior to analysis for the average effect of the medication as estimated from those diagnosed with the conditions but not taking such medications, as described in a prior publication^4^.

### Pathway analysis

Functional enrichment analysis was performed in R using the fgsea^52^ and clusterProfiler^53^ packages for Gene-Set Enrichment Analysis (GSEA). Analysis was performed using pathway annotation from human Reactome (v. 73; 06/2020 release), WikiPathways (07.2020 release) and KEGG (22.07.2020 release) databases. The full SomaScan panel of proteins was used as the background set of proteins. GSEA results were filtered for redundant entries based on semantic similarity between protein groups. Further, UniProt keyword (15.10.2019 release) annotations were used for classification of proteins. Significantly over-represented diseases were assessed using the Genetic Association Database Disease^54^. Clustering in heatmaps was performed using the complete linkage Euclidean distance method.

Voronoi Treemaps were used to visualize absolute beta values from associations between protein changes in conjunction with hierarchical classifications from KEGG Brite Orthology [https://www.genome.jp/kegg/brite.html]^55^. The 3D PCA plot was constructed based on the associations of the protein changes between visits 1 and 4 with all clinical variable changes using z-scored beta values. This was done using the Genedata Analyst v13.0 software (Genedata AG, Basel, Switzerland).

## Data Availability

All data are present in tables in the manuscript and supplementary tables,figures.

## Author contributions

NAY, KS, and SCH designed the study; RE and HS performed the proteomics assays; NAY and SCH performed the data analysis and wrote the manuscript; RE and FS performed the analyses of the protein pathways; TDA, RDM, SCS, and SCH recruited the patients and collected the clinical data and blood; and all authors edited the final manuscript.

The authors declare no competing interests.

## Funding

Supported by a grant (DK-55006) from the National Institute of Diabetes and Digestive and Kidney Diseases of the National Institutes of Health, a U.S. Public Health Service research grant (M01-RR00064) from the National Center for Research Resources, Biomedical Research Program funds from Intermountain Healthcare, and from Qatar Foundation to Weill Cornell Medicine.

## Notes

### Competing Interest Statement

The authors have declared no competing interest.

### Clinical Trial

This is a proteomics study on gastric bypass surgery patients

### Author Declarations

All subjects provided written informed consent and the study was approved by the University of Utah Institutional Review Board.

